# Maximisation of open hospital capacity under shortage of SARS-CoV-2 vaccines

**DOI:** 10.1101/2021.03.08.21253150

**Authors:** Wolfram A. Bosbach, Martin Heinrich, Rainer Kolisch, Christian Heiss

## Abstract

**Motive:** The Covid-19 pandemic has led to the novel situation that hospitals must prioritise staff for a vaccine rollout while there is acute shortage of the vaccine. In spite of the availability of guidelines from state agencies, there is partial confusion about what an optimal rollout plan is. This study investigates effects in a hospital model under different rollout schemes.

**Methods:** A simulation model is implemented in VBA and studied for parameter variation. The implemented code is available as open access supplement.

**Main results:** A rollout scheme assigning vaccine doses to staff primarily by staff’s pathogen exposure maximises the predicted open hospital capacity when compared to a rollout based on hierarchical prioritisation. The effect increases under resource scarcity and increasing disease activity. Nursing staff benefits most from an exposure focused rollout.

**Conclusions:** The model employs SARS-CoV-2 parameters; nonetheless, effects observable in the model are transferable to other infectious diseases. Necessary future prioritisation plans need to consider pathogen characteristics and social factors.

## 1. Introduction

With the availability of Covid-19 vaccinations (1,2), hospital operations management has faced worldwide the new situation that a vaccine rollout scheme for hospital staff had to be implemented. Due to scarcity of vaccine doses, prioritisation decisions have to be made during this rollout. This study makes a contribution to this topic by investigating consequences from different rollout schemes. The study’s focus is the maximisation of open hospital capacity which is assumed to maximise patient benefit.

Research studies from the pre-Corona age exist. Their focus was to prove benefit from vaccination policies (3–5). Shortage of vaccine supply was not a particular focus topic; it was known for e.g. yellow fever (14). Instead rather an oversupply and mandatory vaccination were discussed (6). The early 2020 vaccination framework of the World Health Organisation (WHO) already included aspects of prioritisation decisions for a vaccination rollout against the severe acute respiratory syndrome coronavirus 2 (SARS-CoV-2) (7). The general logistics of the large scale Covid-19 vaccine rollout currently taking place require vaccine doses, vaccinating staff administering the doses, and a process to assign it to patients (8). Rollout recommendations or deployment plans have been developed and published by state agencies (9,10), also with details on the process of vaccine recommendation decisions (11).

There is a general agreement that vaccines should be used to their best potential to curb the pandemic’s consequences, in particular during the early vaccine rollout phase when shortage of vaccine supply prohibits an immediate full rollout to the entire population. Decision making under shortage of resources is known from other important medical supplies in connection with the current pandemic (12,13). Insights into prioritisation decisions and their consequences have been published for studies considering the entire population (15–19). Delaying a 2^nd^ dose if required by drug regime has been discussed as option to reduce vaccine shortage (20,21). In spite of the available material and dedicated rollout recommendations, e.g. (9,10), decision making about rollouts to hospital staff can still prove to be controversial. The weighing in of factors such as hierarchical importance (22,23), or student status (24) can lead to differences in the rollout scheme and prioritisation among hospital staff. The rollout in the United Kingdom so far has been successful (25). Subgroups of a population such as those economically worse off (26), or minorities (27) can be requiring special attention however. Also, nursing staff who by the nature of their work are in close contact to patients must not be forgotten in rollout schemes and prioritisation (28).

In this presented study here, a model hospital is simulated and its capacity is calculated. The hospital’s capacity is limited by available staff. Staff are exposed to the pathogen and are vaccinated after two different rollout schemes which are compared to each other. The first rollout scheme assigns vaccine doses in **hierarchical order top down**. It relates to arguments made in the past (22,23) that higher ranking staff are fewer in number, more important as individuals for the functioning of the hospital, and that they are of greater age which can increase the transmission probability of a pathogen. The alternative rollout scheme first prioritises between hospitals units by **exposure to the pathogen**. Accidents & Emergencies (A&E) where exposure to non-tested outpatients happens is assigned vaccine doses first and hospital wards follow behind. On each unit, vaccines are assigned in hierarchical order.

The model is implemented as Visual Basic for Application (VBA) macro. The macro and its embedded version in Microsoft-Excel are available as open access supplement 1 and 2 under the GNU General Public License version 3, or any later version. We hope that the open access macro code will further exchange and make accessibility of the study’s work easier. A similar project with results for an entire population, split by age groups, is published under (18), pre-corona work about hospital staff under (3,4,6). The study’s methodology uses an algorithm similar to known work from hospital capacity planning (29). Staff and vaccines are modelled as streams, similar as in (30). It applies novel insights into the epidemiological spreading of SARS-CoV-2 (31,32), and factors influencing disease spreading such as age (33,34).

The study’s parameters are set to values typical for the current SARS-CoV-2. However, the methodology and the implemented model are equally applicable to other infectious diseases. Although mostly gone unnoticed in Europe and North America, the list of major infectious disease outbreaks in e.g. Hong Kong during the 25 years before the ongoing SARS-CoV-2 pandemic included avian flu H5N1 in 1997, SARS in 2003, swine flu 2009, and avian flu H7N9 in 2013 (35). Hence, a vaccine rollout in the general population and amongst hospital staff against an infectious disease might be needed again in the future.

## 2. Methodology and model implementation

For the present study, a hypothetical hospital is modelled.

### 2.1. Hospital structure and staff reserve

The hospital is structured in 4 units. Staff is coming to work every day and is simulated for a variation of initially available staff reserve.

#### Hospital staff

Hospital staff consists of doctors and nurses. Nurses are not substructured further. Doctors are each assigned to the rank of physician, senior doctor, executive senior, or chief. The model assumes that any staff member is qualified to work on any hospital unit. Table a summarises the staff numbers initially available at *t* = 0 for the simulated base case and the scenarios of staff reserve variation.

**Table a:** Overview of staffing scenarios

|  | **Base staffing** |  | **Reduced staffing** |  | **Increased staffing** |  |
| --- | --- | --- | --- | --- | --- | --- |
| Rank | Total | Reserve | Total | Reserve | Total | Reserve |
| Chief | 1 | Combined 2 | 1 | Combined 0 | 1 | Combined 4 |
| Executive Senior | 5 |  | 3 |  | 7 |  |
| Senior Doctor | 6 | 2 | 5 | 1 | 8 | 4 |
| Physician | 14 | 4 | 12 | 2 | 18 | 8 |
| Nurse | 33 | 8 | 29 | 4 | 41 | 16 |

#### Hospital units

The hospital is structured in 4 units. It is assumed that the hospital operates under the speciality of general internal medicine. A&E receives out-patients. In-patients are treated on wards 1-3. The daily numbers of staff required for 100%/50% operations of each unit are shown in Table b. Nurses and physicians are required to be present. The chief, executive seniors, and senior doctors provide background service. Chief and executive seniors are expected during staff shortage also to work in the role of senior doctor, they are downward compatible.

**Table b:** Staff requirement for full or partial unit operations

|  | A&E |  | Ward 1 to 3 |  |
| --- | --- | --- | --- | --- |
| Rank | 100% | 50% | 100% | 50% |
| Chief | Combined 1 | Combined >0 | Combined 1 | Combined >0 |
| Executive Senior |  |  |  |  |
| Senior Doctor | 1 |  | 1 |  |
| Physician | 4 | >1 | 2 | >0 |
| Nurse | 10 | >4 | 5 | >2 |

Hospital capacity Γ is calculated as average of the open status of A&E, and ward 1-3 in Eq. 1.

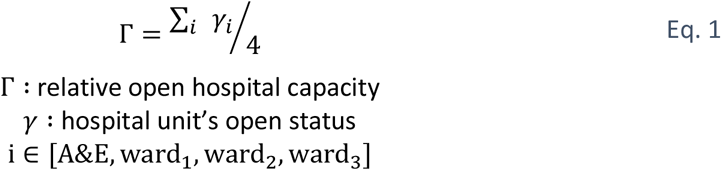

### 2.2. Age distribution

It is known that there can be an age factor in the epidemiology of a pathogen. SARS-CoV-2 is no exception in that regard (33,34). For each staff member, we assign an age based on a uniform distribution. Table c gives the bounds of the distribution for the different staff roles. The impact of age on infection events is discussed further in Section 2.4.

**Table c:** Lower bound and upper bound for age distribution

| Rank | Lower bound | Upper bound |
| --- | --- | --- |
| Chief | 51 | 65 |
| Executive Senior | 41 | 50 |
| Senior Doctor | 33 | 40 |
| Physician | 25 | 32 |
| Nurse | 18 | 65 |

### 2.3. Vaccination rollout

The model compares two rollout schemes. In each case, the rollout rate *v*_*S*_ defines the number of vaccines per day available to the hospital. Staff are vaccinated, unless they are symptomatic, or already vaccinated. The model is implemented for a vaccine which applies only 1 dose per person.

#### Top down vaccine rollout

In the case of the top down rollout scheme, the model assigns doses hierarchically and moves top down, starting with the chief. Senior executives, senior doctors, and physicians follow. Nurses are assigned doses last.

#### Exposure focused vaccine rollout

The exposure focused rollout scheme prioritises staff by their exposure to the pathogen during their hospital shifts. Due to symptomatic and non-symptomatic out-patients walking in at A&E, A&E staff’s exposure is assumed greater than exposure of staff on the wards 1-3. A&E staff are vaccinated first; staff on wards 1-3 follow unit by unit in chronological order. On A&E and wards 1-3, again a hierarchical vaccine assignment is implemented.

### 2.4. Disease status and pathogen transmission

The model calculates each day for each staff member a pathogen transmission probability and the individual disease status.

#### 2.4.1. Staff member’s disease status

The disease status of a staff member can be non-infected, infected and non-symptomatic, or infected and symptomatic. After pathogen transmission, a staff member is infected, non-symptomatic and continues working. After the passing of the incubation time *t*_*inc*_ (36), the staff member becomes symptomatic and remains off duty for the recovery duration of *t*_*rec*_. Whether reinfections with SARS-CoV-2 are possible and by what probability is being investigated at the moment (37). The model assumes that staff can reinfect unless vaccinated. For simplification of disease behaviour, staff being vaccinated during *t*_*inc*_ is assumed again not-infected.

#### 2.4.2. Pathogen transmission

The model assumes that the pathogen transmits from infected human to non-infected human. The model assumes that vaccinated staff are immune to the pathogen and are not infectious for other staff members. For each working day, a probability Π is calculated for which a staff member’s status is changed to infected. This probability considers the level of exposure, *R*_*S*_ as used in epidemiology as reproductive number (31), the age (33,34), and the reduced patient contact of greater hierarchy ranks. Eq. 2 and Eq. 3 below detail the calculation of the probabilities Π_*A&E*_ and Π_*ward*1−3_.

##### Pathogen exposure on A&E and wards 1-3

Exposure quantifies in the model the contact to infected humans. On A&E, staff comes into contact with out-patients who walk in and are positive with probability rate *p*_*pos*_. The model assumes that on A&E each staff member has contact with 20 patients during one shift. The model adds the number of infected, non-symptomatic colleagues 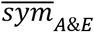 on A&E who come to work on that day and increase pathogen exposure to other staff members.

On wards 1-3, outside A&E, the model assumes that all patients are tested and if positive isolated so that patients cause no pathogen exposure to staff. The model adds again the number of infected, non-symptomatic colleagues 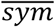 who come to work on that day to ward 1-3. A base probability of 1/100 is added. By this, the model accounts for the fact that pathogen intake on wards 1-3 vanishes compared to intake on A&E but that it is not nil.

##### Infection risk Π due to exposure

Infection risk due to exposure is modelled according to research insights about reproduction numbers found in (31,32) for SARS-CoV-2. These studies have provided probabilities by which an individual can expect to be infected after a specified exposure event (specified by parameters such as pathogen emission rate, breathing activity, aerosol concentration, or duration of exposure). In this context, the parameter *R*_*S*_ (pathogen characteristic reproduction number) is used. *R*_*S*_ is defined as the number of people infected by one index patient. This model now uses the values for *R*_*S*_ of (31), multiplies by the number of positive contacts, and divides it by factor 5 for obtaining the staff member’s personal risk Π. The reduction by factor 5 is based on the assumption that the relevant index patient has contact to 5 members of staff.

##### Amendment for age and reduced patient contact of greater hierarchy ranks

It is known that social status and age can influence the spreading of infectious diseases (24,26,27). The model acknowledges this and considers two additional amendment factors which are both multiplied on the infection risk Π.

The age factor *T*_*S*_ is a number greater or equal 1 and scales linearly between the staff age of 18 and 65, assigned for each staff in Section 2.2. Values are set to the magnitude as known so far for SARS-CoV-2 (33,34).

In modern hospitals, higher hierarchy ranks have typically less patient contact compared to junior doctors or nurses. The model considers this by a factor *H*_*S*_ which is set to a value greater or equal 1. The infection risk for the hospital’s chief and executive seniors is divided by *H*_*S*_.

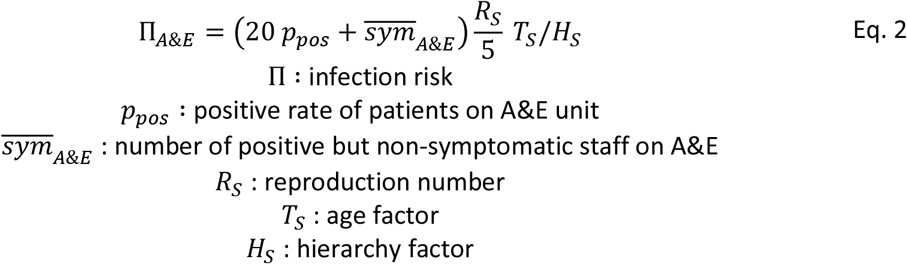

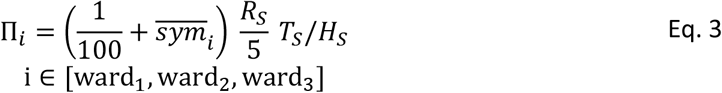

### 2.5. Assignment of staff to units and open/closed-status definition

Staff who is not symptomatic is assigned to work on the hospital’s units. Available staff is by first priority assigned to A&E; followed by wards in ascending ward number. Symptomatic staff is considered off duty. Open/closed status of a unit is defined for each day by the model according to staff requirements of Table b.

### 2.6. VBA implementation

The model has been implemented in VBA. It calculates for a period of 100 days the disease spreading under different scenarios. Each scenario is run *n* = 500 times to account for statistical unevenness of random number function of e.g. age distribution (Section 2.2) or pathogen transmission (Section 2.4.2). The VBA macro is run for this present study in a Microsoft-Excel environment on an Intel(R) Core(TM) i5-6200U CPU @ 2.30GHz with 8.00 GB memory RAM. Computation time for 500 cycles of each scenario lies at around 25 minutes, increasing/decreasing with increasing/decreasing staff size.

## 3. Results and discussion

The VBA implementation of the hospital model is run for different scenarios of parameter values. The meaningfulness of the study’s results lies not in individual values but the trends which can be observed for parameter variation. Disease parameters or hospital parameters might change for future scenarios. The found effects are valid also for other future settings.

### 3.1. Base case and statistical convergence

Table d shows the model parameters and their values in the base case. The effect of the parameters on e.g. open hospital capacity Γ is discussed in relation to this base case.

**Table d:** Parameters of the base case scenario

| Parameter | $v_S$ [1/d] | $R_S$ [-] | $p_{pos}$ [-] | $t_{inc}$ [d] | $t_{rec}$ [d] | $T_S$ [-] | $H_S$ [-] | Staffing |
| --- | --- | --- | --- | --- | --- | --- | --- | --- |
| Value | 1 | 7 | 0.25 | 5 | 10 | 2.5 | 5 | Base staffing |

Figure 1 shows the output of the model for predicted open hospital capacity, averaged for *n* = 500 runs together with its standard deviation per day for the duration *t* of 100 days. The figure shows that under the top down rollout scheme (blue) a greater decrease in open hospital capacity is expected than under the exposure focused rollout (green). In both cases, the minimum capacity is reached after passing of incubation time and recovery time (15 days). In the beginning, expected open capacity decreases over time as non-vaccinated staff contract the pathogen. This fall does not set in at *t* = 1 day as staff reserves are still available initially. After incubation time and recovery time, the first staff members who contracted the pathogen return to work. The ensuing increase of open hospital capacity is the result of greater immunity of staff to the pathogen due to the vaccine rollout. The fall to the minimum capacity is steeper for the top down rollout, and the following return to full capacity after is more moderate for the top down rollout when compared to the green exposure focused rollout.

**Figure 1:**
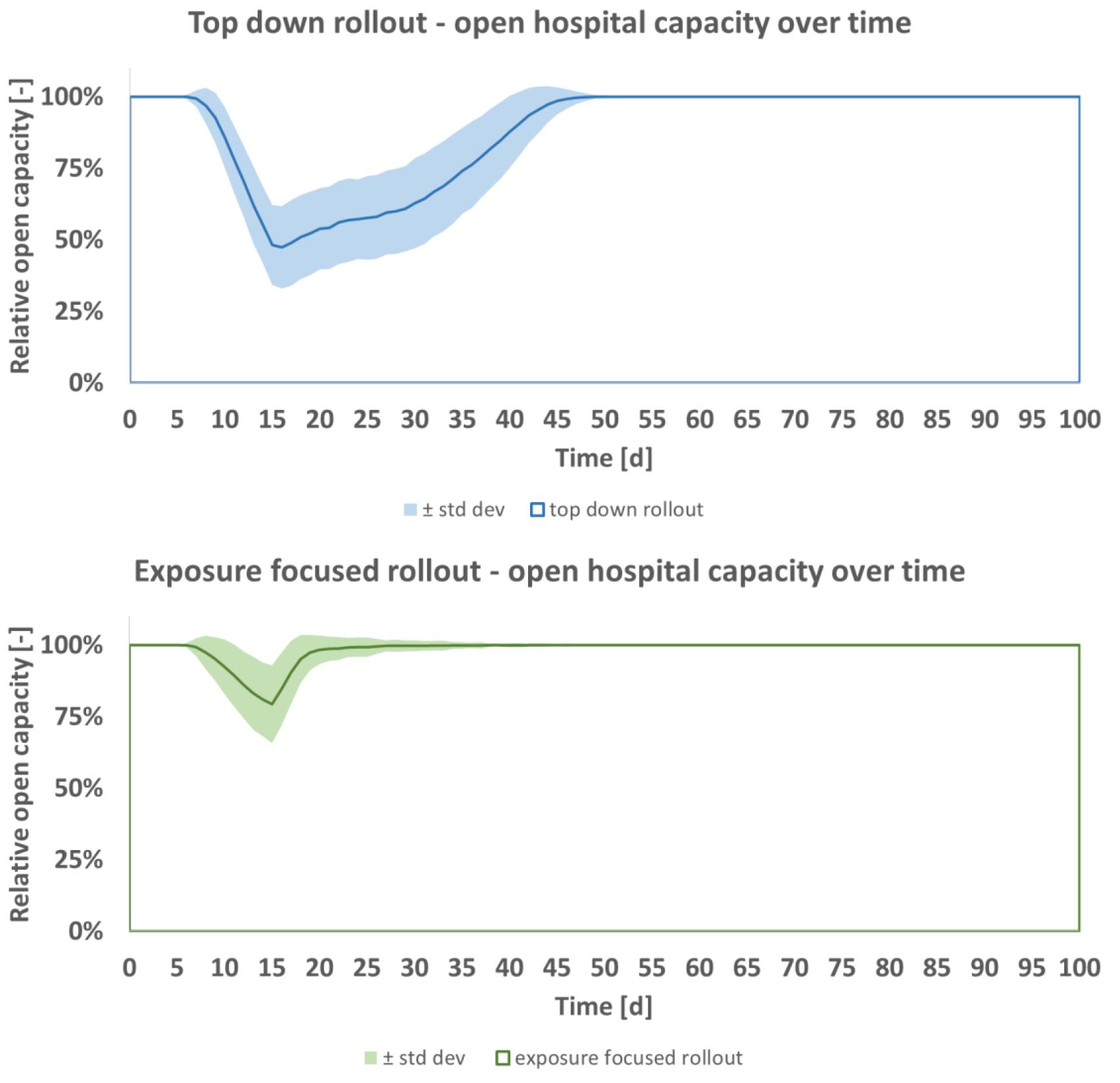
Open hospital capacity, predicted mean ± standard deviation per day, for top down (blue) and exposure focused (green) vaccine rollout, base case scenario of Table d

Figure 2 shows for predicted hospital capacity in the base case of Table d the average and the average’s standard deviation per *n* as developing over *n*. Both rollout schemes converge towards a constant after around *n* = 100. Standard deviation lies below 0.4% and is greater for the top down vaccine rollout (blue). For both rollout schemes, standard deviation converges towards a constant value and changes only negligibly after *n* = 300.

**Figure 2:**
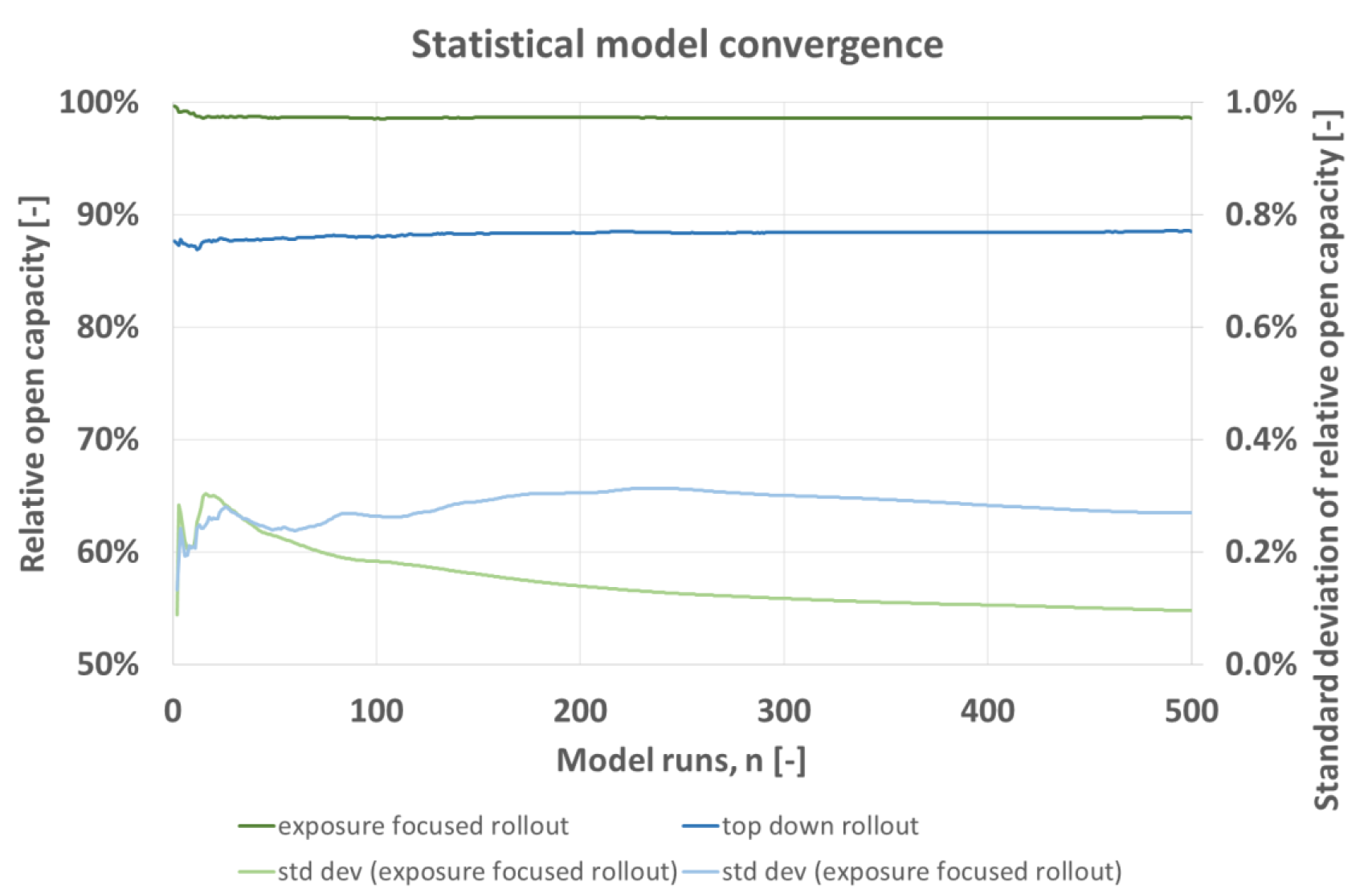
Statistical model convergence, expected relative open capacity averaged for n runs with corresponding standard deviation per model run n, for top down (blue) and exposure focused (green) vaccine rollout, base case scenario of Table d

### 3.2. Disease activity: Influence of pathogen infectiousness and prevalence

Prevalence *p*_*pos*_ of the disease in out-patients coming into A&E is the main variable determining pathogen intake into the hospital. Infectiousness *R*_*S*_ decides about the number of transmission events from patients onto staff, or between staff. To investigate the effect of increased disease spreading, a parameter study is performed where the parameters *R*_*S*_ and *p*_*pos*_ are varied according to the values given in Table e while all other parameters are kept as described in the model’s base case scenario of Table d. *R*_*S*_ is in the magnitude as obtained in (31) for Sars-Cov-2. *p*_*pos*_ is varied between 0.1 and 1.

**Table e:**
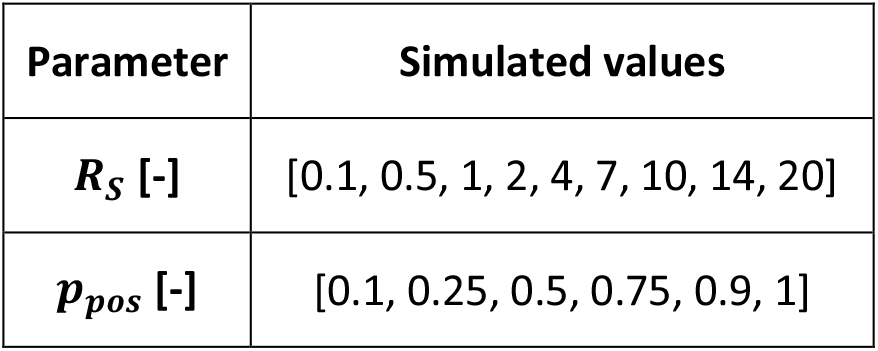
variation of infectiousness and exposure

| Parameter | Simulated values |
| --- | --- |
| $R_S$ [-] | [0.1, 0.5, 1, 2, 4, 7, 10, 14, 20] |
| $p_{pos}$ [-] | [0.1, 0.25, 0.5, 0.75, 0.9, 1] |

Figure 3 shows the capacity for both rollout schemes for all resulting *p*_*pos*_ ∗ *R*_*S*_. The model predicts that for greater disease activity the advantage of the exposure focused rollout increases until a nearly constant level is reached at around *p*_*pos*_ ∗ *R*_*S*_ = 5. At this constant level, predicted open capacity under the exposure focused rollout is about 1.2 times greater than what is predicted for the top down rollout (orange). When disease activity decreases, the advantage of the exposure focused rollout does so as well. Both rollout schemes achieve approximately equal results for 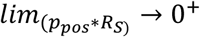.

**Figure 3:**
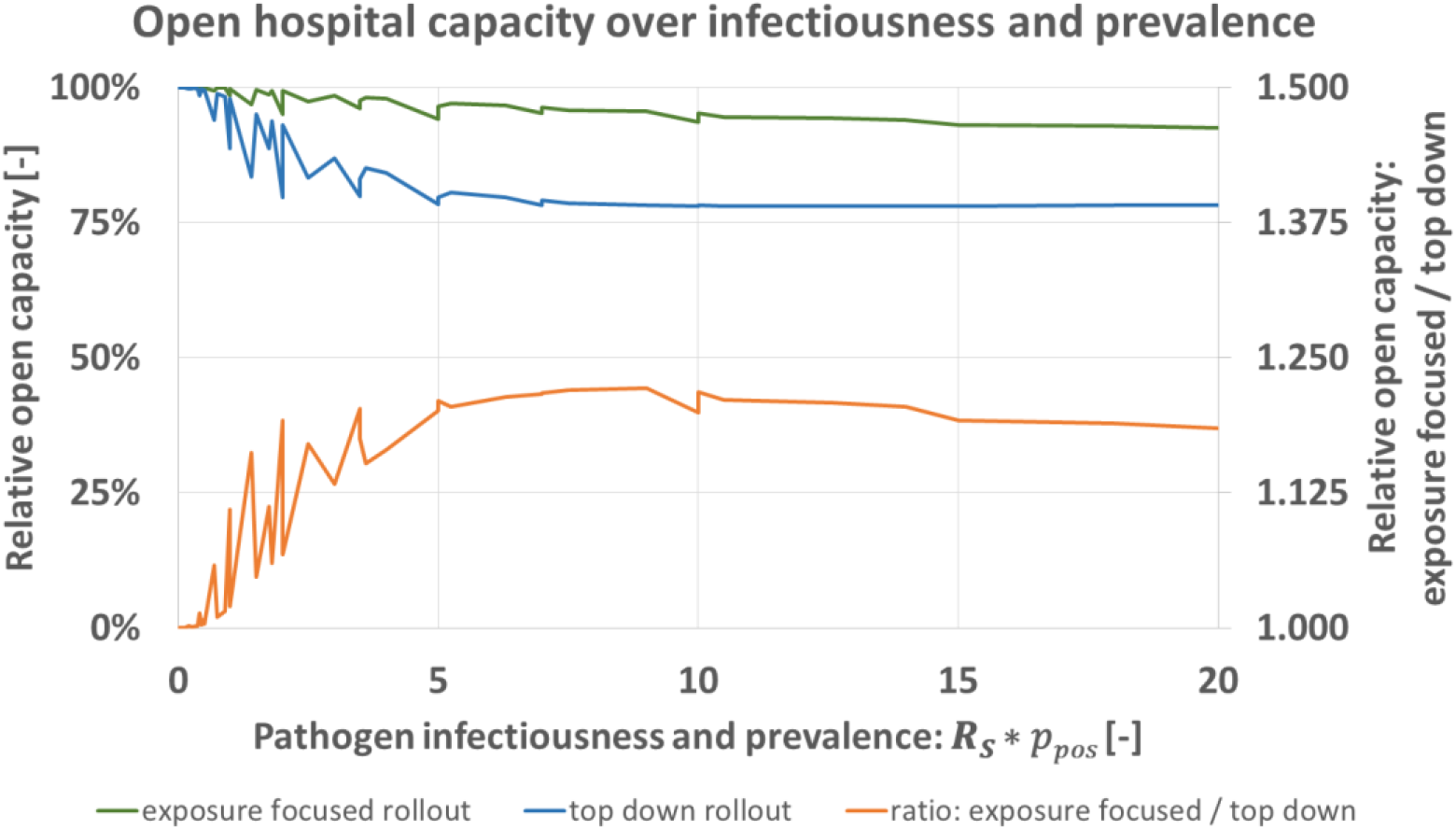
Predicted open hospital capacity over product of infectiousness R_S_ and prevalence in A&E-patients p_pos_ for top down (blue), and exposure focused rollout (green), together with their respective ratio (orange)

Figure 4 gives information about the influence of disease activity on open status on unit level. The base case scenario (*R*_*S*_ = 7, *p*_*pos*_ = 0.25, Table d) leads to a temporary total closure of ward-3 and substantial closures of ward-2 and ward-1 under the top down rollout. The prediction for exposure focused vaccine rollout of the base case predicted a fully open ward-1. Ward-2 and 3 would be expected to lose temporarily capacity, by a margin less than what is predicted for the top down scheme. If disease activity is increased to e.g. (*R*_*S*_ = 10, *p*_*pos*_ = 0.75) under both rollout schemes open capacity drops. Temporarily ward-1 to 3 are predicted closed and A&E predicted not fully open under the top down rollout while A&E is predicted fully open under the exposure focused rollout.

**Figure 4:**
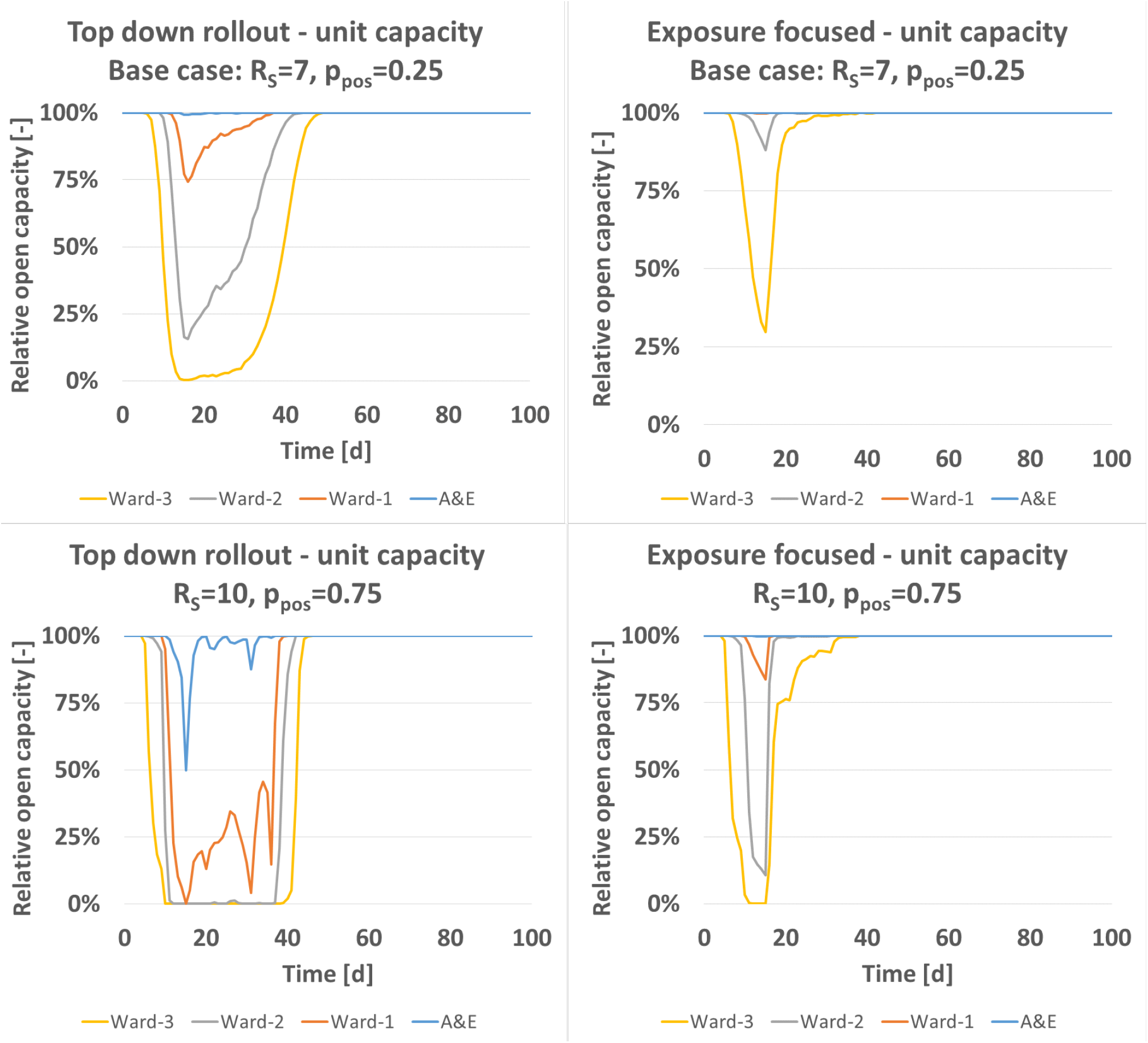
Relative open capacity split onto unit level for base case (R_S_ = 7, p_pos_ = 0.25, Table d), and parameter variation (R_S_ = 10, p_pos_ = 0.75)

### 3.3. Influence of recovery time and social considerations

The time needed by staff to recover *t*_*rec*_ [d] from the simulated pathogen can vary, based on the pathogen and predisposition of a staff member. To quantify the influence on the model’s prediction, *t*_*rec*_ is simulated for the values shown in Table f while setting all other parameters to the base case of Table d. Figure 5 shows the obtained results, formatted as before Figure 3 (exposure focused rollout green, top down rollout blue, ratio of both schemes orange).

**Table f:** variation of recovery time

| Parameter | Simulated values |
| --- | --- |
| $t_{rec}$ [d] | [1, 5, 10, 14, 20] |

**Figure 5:**
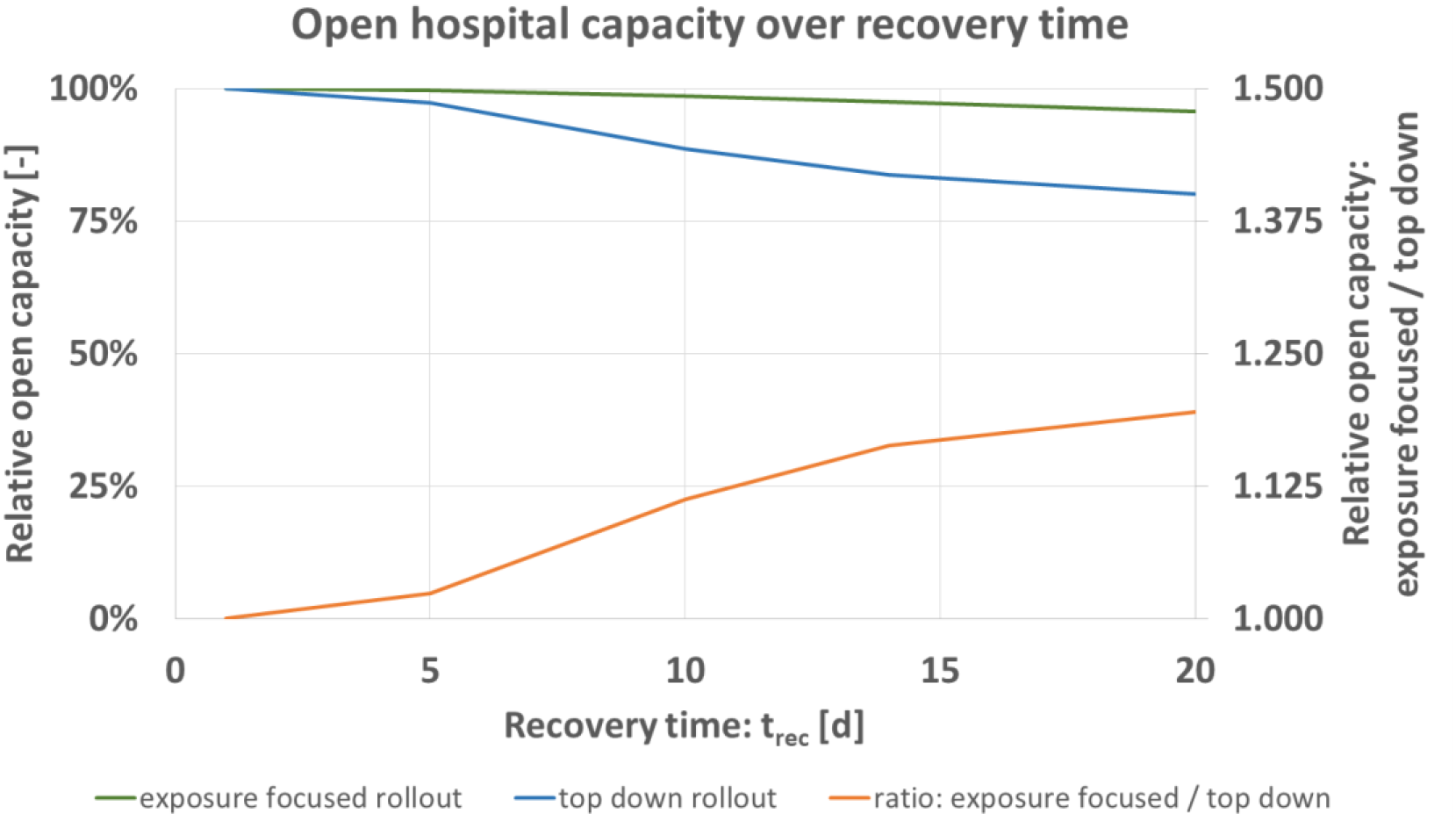
Predicted open hospital capacity over recovery time t_rec_ [d] for top down (blue), and exposure focused rollout (green), together with their respective ratio (orange)

Figure 5 shows that expected open hospital capacity decreases for greater *t*_*rec*_ of infected staff. The advantage of the exposure focused rollout increases for greater *t*_*rec*_. Similarly to Figure 3 for disease activity, the difference factor (orange) lies at around 1.2 for the maximum value simulated.

Figure 6 gives the expected number of infected staff over time, top down rollout scheme subtracted from exposure focused rollout. Total numbers are shown by hierarchy group. This analysis is of importance as it relates to work which showed that social factors must be considered during the current pandemic (24,26,27). The greatest difference between the vaccine rollout schemes is seen for the nursing staff. Their infection numbers are predicted by the model to increase most, once the hierarchical vaccine allocation takes place. Executive seniors and senior doctors are predicted to in average benefit from the top down rollout. The chief who also can work on A&E is vaccinated fairly early in both schemes and not predicted to contract the pathogen. Interestingly, the physicians are treated favourably in each of the two rollout schemes. Depending on progression of the rollout they benefit or not. Initially, they are worse of as doctors of greater hierarchical rank are assigned vaccine doses. After *t* of 27 days, their group however benefits from the top down scheme as they are assigned vaccine doses which would go alternatively to A&E nurses under the exposure focused rollout.

**Figure 6:**
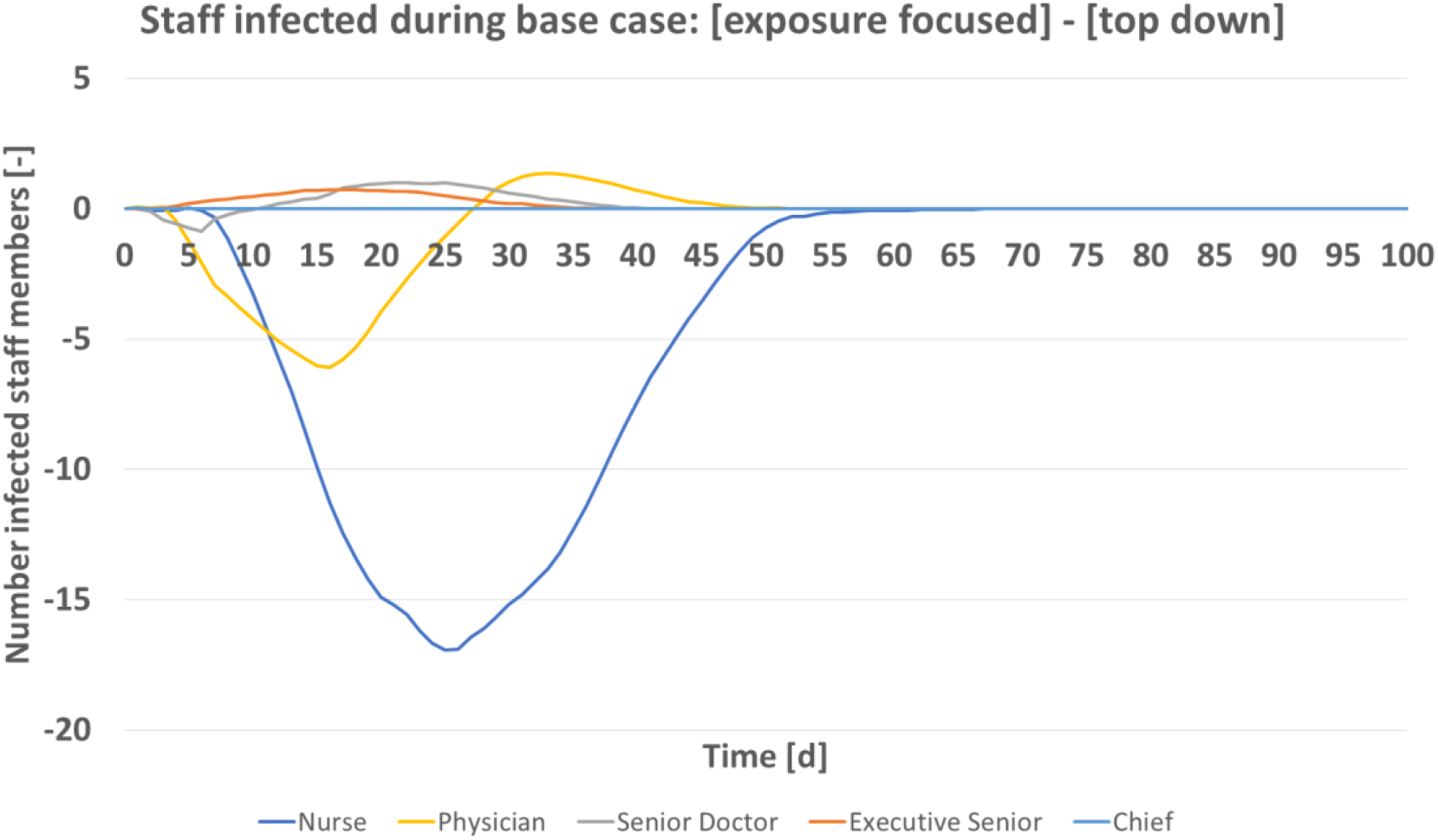
Predicted number of infected staff by hierarchy group (Table a) over time for base case scenario (Table d)

### 3.4. Availability of vaccine and rollout rate

A very urgent aspect of the current rollout is the scarcity of vaccine doses as a resource. Different vaccines have been approved by regulatory bodies, more are in the final stage of certification (1,2). However, production numbers do not yet provide sufficient supply for an immediate full rollout. In today’s real world situation, vaccines are rationed (25). The model predicts for the two rollout schemes that the difference in expected open hospital capacity increases for greater resource scarcity, Figure 7 and Table g.

**Table g:** variation of daily rate of vaccines

| Parameter | Simulated values |
| --- | --- |
| $v_s$ [1/d] | [0, 1, 2, 5, 7, 10, 20, 30, 40] |

**Figure 7:**
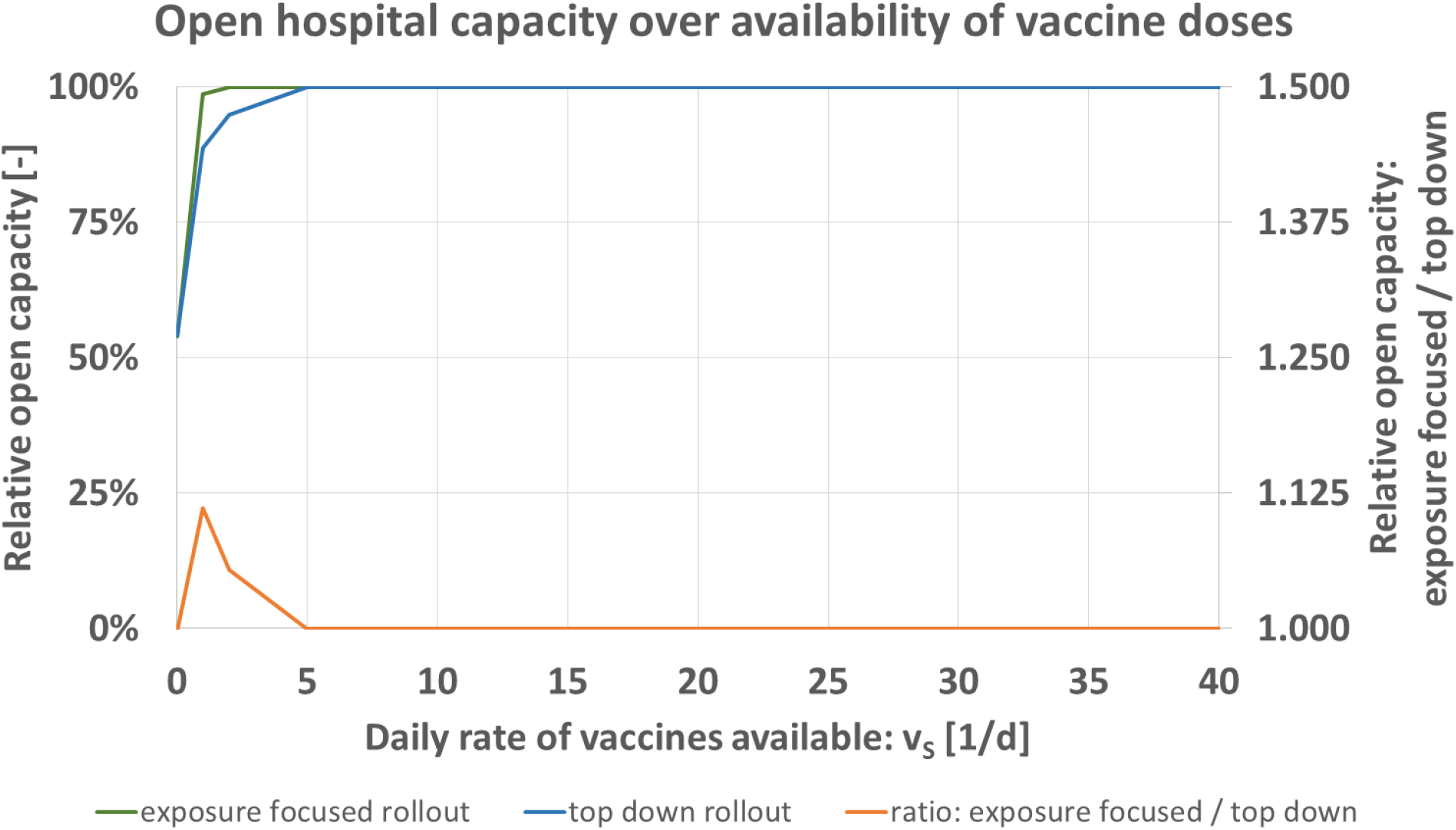
Predicted open hospital capacity over daily rate of vaccines available v_S_ [1/d] for top down (blue), and exposure focused rollout (green), together with their respective ratio (orange)

The availability of vaccine doses *v*_*S*_ [1/d] is iterated for the values of Table g while keeping all other parameters as specified for the base case Table d. Figure 7 shows that the maximum of the relative comparison between both schemes (orange) is located at 1/d. For the staff size as specified in Table a, no measurable difference exists in the model once supply of *v*_*S*_ >5/d is available. This means that in particular in situations of resource scarcity, the exposure focused rollout scheme is advantageous when measured in open hospital capacity.

### 3.5. Influence of staff reserve

Another resource in hospital operations apart from the vaccines per day available for the hospital is the staff. The model calculates a partial or full closure of units as penalty condition for staff shortage due to disease, Table b and Table h. For the variation of initial staff reserve, the model predicts that in each scenario the exposure focused rollout scheme leads to greater expected open hospital capacity.

Figure 8 shows the numbers obtained for the variation of staff reserve. Similarly as before in Figure 7 for the availability of vaccine doses, the difference obtained for exposure focused and top down rollout is greater when base staffing or only reduced staffing is initially available. With increased staffing, the difference between the two rollout schemes decreases. Here again, results can be interpreted in a way that under shortage of resources the exposure focused vaccine rollout is more vital for the hospital’s ability to operate.

**Figure 8:**
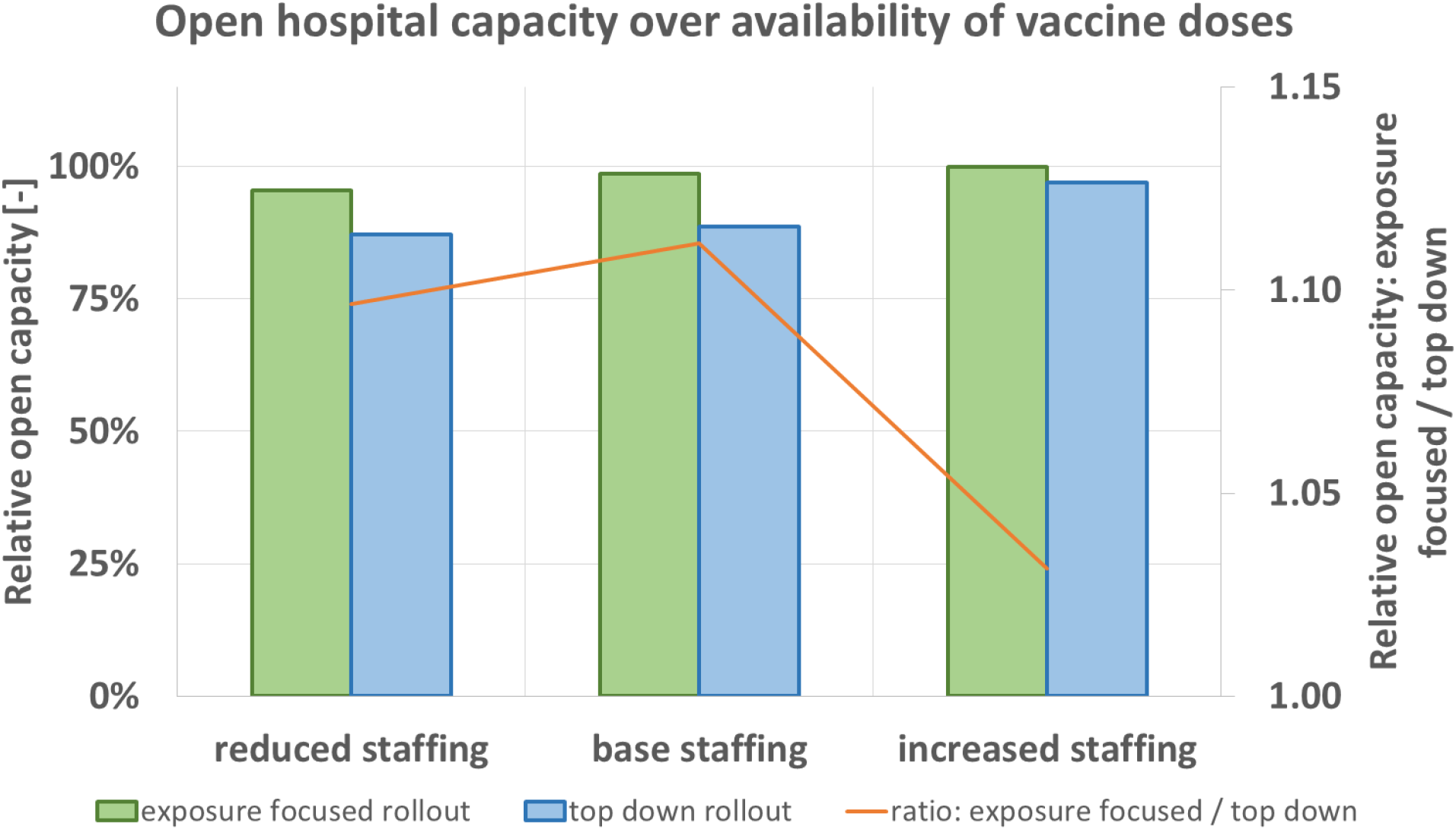
Predicted open hospital capacity depending on initial staff reserve (Table a) for top down (blue), and exposure focused rollout (green), together with their respective ratio (orange)

**Table h:** variation of staff size and initial staff reserve, numbers defined in Table a

| Parameter | Simulated values |
| --- | --- |
| Staffing | [Base staffing, Reduced staffing, Increased staffing] |

### 3.6. Consideration of increased transmission due to age and roles in hospital hierarchy

In the argumentation for the case of the top down rollout scheme, the special importance of higher staff ranks for hospital operations is one factor. On top, their typically greater age makes them more susceptible for pathogen transmission. Their exposure to patients is typically reduced compared to nurses and junior doctors. Higher ranks are involved in administrative tasks which during the pandemic can partially even be done in home office. The model is iterated over *T*_*S*_ [-] (increased pathogen transmission probability for greater age) and *H*_*S*_ [-] (reduced patient contact for the hospital’s chief and executive seniors) to demonstrate the influence of both variables. Values are shown in Table i. Both are varied between 1 and 20.

**Table i:** variation of age factor and reduced exposure due to hierarchy

| Parameter | Simulated values |
| --- | --- |
| $T_S$ [-] | [1, 1.5, 2.5, 5, 10, 20] |
| $H_S$ [-] | [1, 2, 5, 10, 20] |

Figure 9 shows the predicted open hospital capacity of the base case (Table d) for the variation of age dependency of pathogen transmission probability, T_S_. Under both rollout schemes, expected open capacity decreases for greater age factor. The model predicts better outcome for the exposure focused rollout (orange). This means that the influence of age (distribution per hierarchy group in Table c) does not serve as an argument for the case of the top down rollout scheme but rather against it. It must not be forgotten that the age factor applies to all hospital staff. Nurses realistically also reach age of 60 and over as the hospital’s chief when they are equally more probable to contract the pathogen.

**Figure 9:**
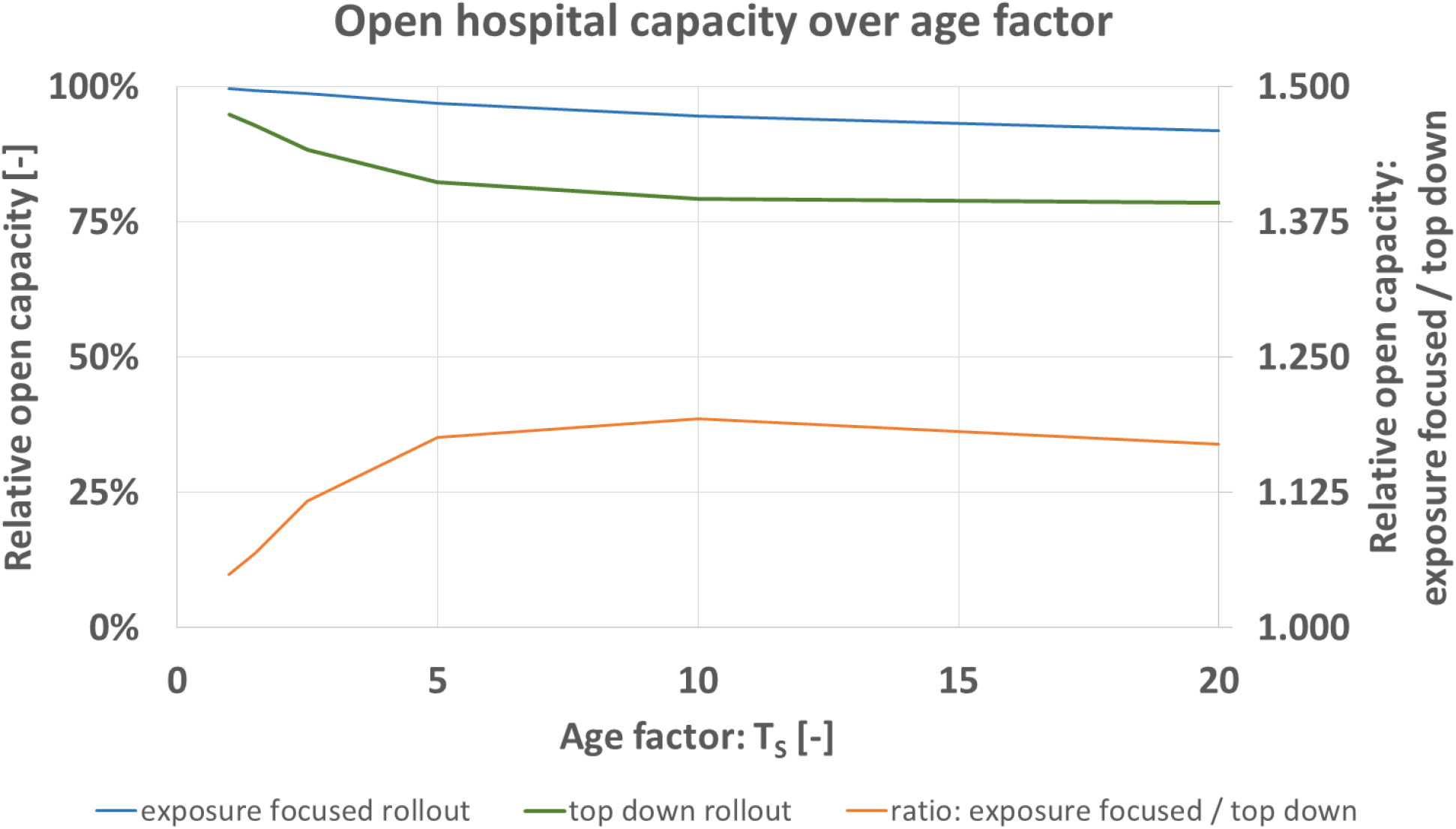
Predicted open hospital capacity over age factor T_S_ [-] for top down (blue), and exposure focused rollout (green), together with their respective ratio (orange)

The influence of *H*_*S*_ is found interestingly to be negligible in the hospital model as defined in the present study. No clear influence on expected open hospital capacity or on infection numbers amongst the chief and executive seniors is predicted in the per cent or per mill range. Increasing the number of statistical cycles to cycle numbers greater than the applied *n* = 500 is possible. It isn’t performed as part of the present study however as it would generate numerical results which suggest pseudo precision that is beyond the predictive precision of the hospital model.

This lack of explanatory power of the hospital model with regard to *H*_*S*_ is explicable by the rollout schemes as defined under Section 2.3. The chief and executive seniors are first to be vaccinated for the top down scheme. In the exposure focused rollout scheme, the chief or the executive senior on A&E shift is the first person of the entire staff to be vaccinated at *t* = 1 day. Under this setting, exposure to the non-vaccinated chief or executive seniors is minimal already and the influence of *H*_*S*_ does not reduce substantially further the probability of pathogen transmission.

## 4. Conclusions and future work

The model’s implementation in VBA is iterated for parameter studies. The two rollout schemes for a top down or exposure focused assignment of vaccine doses to staff can be compared in various settings. In this hospital model, the expected open hospital capacity which is assumed to maximise patient benefit is generally greater for the exposure focused rollout. The model relies on the drawing of random numbers for e.g. age and transmission events. Building statistical averages for *n* = 500 model runs ensures convergence of results with a constant, Figure 2. Standard deviation of open hospital capacity calculated over averages of *n* lies below 0.4%.

Results predict an increasing advantage by the exposure focused vaccine rollout scheme under increasing disease activity defined by greater pathogen infectiousness and prevalence in patients, Figure 3. Overall open hospital capacity can be broken down in open status of hospital units (A&E, ward-1 to 3). Trends observable for total capacity translate into effects on unit level. Following unit prioritisation, predicted loss of unit open status is more pronounced under the top down rollout scheme. Also an increasing recovery time leads to greater advantage of the exposure focused rollout, Figure 5.

The results in Figure 6 demonstrate the different infection numbers per hierarchy groups. The top down rollout scheme would be most disadvantageous for nurses. Their infection numbers increase greatly under that scheme. Also nurses on A&E get vaccinated only after all doctors have been assigned doses. These comparisons between prioritisation schemes are of importance. It is known that social status, or minority status influences disease spreading (24,26,27).

Shortage of resources (supply of vaccine doses, or initial staff reserve) decreases the predicted open hospital capacity. The exposure focused rollout scheme leads to better results in those scenarios and is the preferable option when compared to the top down rollout scheme, Figure 7 and Figure 8. The influence of greater pathogen transmission probability by greater age is another argument for the case of the exposure focused rollout scheme. Here again, the role of nursing staff who realistically reach ages of 60 and over must be considered.

The implemented and presented model is a first step to investigate decision making strategies for a novel problem (22,23). The model’s complexity is kept to a minimum. Several other ongoing hospital processes could be implemented in it. Extensions could investigate effects of staff moving between hospitals, only partial immunity of staff after a 1^st^ vaccine dose, or the fact that tests produce false negative and false positive results. Also the organisational structure of the hospital could be extended so that it contains several medical specialities; surgery, medicine, radiology, and/or anaesthesia. For a hospital’s ability to perform pre-surgical imaging diagnostics and surgery itself, staffing in radiology and in anaesthesia is essential.

The mathematical evaluation in the presented model implementation builds averages over time. In a more complex implementation of a more diversified hospital model, integrating open status over time and comparison of areas (closed/open) can increase explanatory power.

The model so far only quantifies impact on unit open status. This could be extended into calculations about number of procedures and number of patients affected. The number of patients treated and remuneration by diagnosis-related groups would allow a quantification of the financial impact on annual business results of the hospital.

The scientific understanding of SARS-CoV-2 behaviour is expected to increase. Based on this, SARS-Cov-2 mutants, and other future scenarios, new vaccine rollouts might become necessary. After 4 major outbreaks of infectious diseases in e.g. Hong Kong during the 25 years before the Corona pandemic alone (35), similar situations might repeat in the future.

Deposited at doi.org/10.5281/zenodo.4589332

**Supplement-1**. << macro.vba>>

**Supplement-2**. << macro_in_excel.xlsm>>

## Data Availability

The model is implemented as Visual Basic for Application (VBA) macro. The macro and its embedded version in Microsoft-Excel are available as open access supplement 1 and 2 under the GNU General Public License version 3, or any later version (doi.org/10.5281/zenodo.4589332). We hope that the open access macro code will further exchange and make accessibility of the study's work easier.

## Acknowledgements

The authors thank Prof Pawel Dlotko from the Dioscuri-Centre for TDA in Warsaw (Poland) for his advice on open access software licensing. The Deutsche Forschungsgemeinschaft (DFG) provided mobility funds through grant BO 4961/6-1. The DFG had no role in writing-up or decision where to submit.

